# A preliminary attempt to harmonize using physics-constrained deep neural networks for multisite and multiscanner MRI datasets (PhyCHarm)

**DOI:** 10.1101/2025.02.07.25321867

**Authors:** Gawon Lee, Dong Hye Ye, Se-Hong Oh

**Author notes:** Correspondence to: Se-Hong Oh, PhD, Department of Biomedical Engineering, Hankuk University of Foreign Studies, 81, Oedae-ro, Yongin, 17035, Republic of Korea., Dong Hye Ye, PhD, Department of Computer Science, Georgia State University, 33 Gilmer Street SE, Atlanta, GA 30303, Georgia, USA. Se-Hong Oh and Dong Hye Ye contributed equally to this work.

## Abstract

In magnetic resonance imaging (MRI), variations in scan parameters and scanner specifications can result in differences in image appearance. To minimize these differences, harmonization in MRI has been suggested as a crucial image processing technique. In this study, we developed an MR physics-based harmonization framework, Physics-Constrained Deep Neural Network for Multisite and multiscanner Harmonization (PhyCHarm). PhyCHarm includes two deep neural networks: (1) the Quantitative Maps Generator to generate T_1_- and M_0_-maps and (2) the Harmonization Network. We used an open dataset consisting of 3T MP2RAGE images from 50 healthy individuals for the Quantitative Maps Generator and a traveling dataset consisting of 3T T_1_w images from 9 healthy individuals for the Harmonization Network. PhyCHarm was evaluated using the structural similarity index measure (SSIM), peak signal-to-noise ratio (PSNR), and normalized-root-mean square error (NRMSE) for the Quantitative Maps Generator, and using SSIM, PSNR, and volumetric analysis for the Harmonization network, respectively. PhyCHarm demonstrated increased SSIM and PSNR, the highest Dice score in the FSL FAST segmentation results for gray and white matter compared to U-Net, Pix2Pix, and CALAMITI. PhyCHarm showed a greater reduction in volume differences after harmonization for gray and white matter than U-Net, Pix2Pix, or CALAMITI. As an initial step toward developing advanced harmonization techniques, we investigated the applicability of physics-based constraints within a supervised training strategy. The proposed physics constraints could be integrated with unsupervised methods, paving the way for more sophisticated harmonization qualities.

## 1. INTRODUCTION

In magnetic resonance imaging (MRI) research, differences in the scanners used may lead to differences in the imaging findings, a phenomenon known as the scanner effect. For instance, several multisite studies of multiple sclerosis have demonstrated variations in the size of lesions when different scanners are used (Biberacher et al., 2016; Sampat et al., 2010; Shinohara et al., 2017). Other studies have reported differences in quantitative values (T_1_ or T_2_) despite the expectation that there would be no variations in these data (Bane et al., 2018 ; Hanson et al., 2020 ; Keenan et al., 2021 ; Stikov et al., 2015). Methods to minimize these discrepancies are therefore needed to standardize results from various scanners.

MR harmonization is an image processing technique that minimizes variations in MR images from different acquisition dates, scanners, or sites. Harmonization methods are grouped into two categories: (1) techniques to standardize data acquisition and (2) techniques to standardize image features through data processing. A recent study involving the standardization of data acquisition presented a methodical approach to reduce variations among different sites by establishing a network that connects MR scanners (Karakuzu et al., 2022). In terms of data processing, two methods can be used: statistical bias elimination and deep neural network–based domain translation. The statistical bias elimination method is commonly performed using conventional methods such as standardization, global scaling, intensity matching, N4 bias correction (Tustison et al., 2010), or the ComBat method (Beer et al., 2020; Choi, 2022; Fortin et al., 2017; Fortin et al., 2018; Garcia-Dias et al., 2020; Pomponio et al., 2020). Although these approaches are accessible to users, they are limited to completely eradicating scanner bias. Additionally, ComBat assumes that the input data follows a normal distribution, which may not always be the case.

For deep learning–based methods, harmonization techniques have been conducted with various network architectures (Bashyam et al., 2022; Fatania et al., 2022; Guan et al., 2021; Ren et al., 2021; Robinson et al., 2020; Yang et al., 2020; Zhang et al., 2019), including U-Net (Ronneberger et al., 2015), Pix2Pix (Isola et al., 2017), GAN (Goodfellow et al., 2020), CycleGAN (Zhu et al., 2017), a variational autoencoder (VAE) (Kingma and Welling, 2013), and StarGAN (Choi et al., 2018). For the U-Net-based method, the DeepHarmony technique employs a 2D U-Net architecture for image-to-image translation (Dewey et al., 2019). Various other harmonization models have been proposed, including the CALAMITI model, which employs the VAE to extract shared anatomical features from two sets of intermodality contrasts, specifically T_1_-weighted (T_1_w) and T_2_-weighted (T_2_w) images (Zuo et al., 2021). A conditional VAE model is also used to harmonize diffusion-weighted MR images for unpaired data (Moyer et al., 2020). However, these models have limitations, such as the loss of structural information during harmonization (Torbati et al., 2023).To mitigate the likelihood of this particular limitation, the MISPEL model employs a supervised harmonization technique using latent representations (Torbati et al., 2023). These methods have the advantage of being less dependent on the target scanner’s images by employing deep neural networks to find common features between two images rather than relying on direct mapping. From an MR physics perspective, quantitative maps, such as T_1_-map, T_2_-map, and M_0_-map, could be regarded as shared features between two MR images when applied to the same subject.

In this study, we propose using the Bloch equation to enhance the harmonization of MR images while maintaining anatomical features. The Bloch equation elucidates the correlation between the MR signal and the magnetic field strength, scan parameters, and quantitative parameter values (T_1_, T_2_, and proton density [M_0_]). Theoretically, by manipulating the scan parameters, the Bloch equation allows one to generate MR image contrasts using quantitative maps. Based on this theoretical assumption, we created a novel approach called PhyCHarm, which is an end-to-end Physics-Constrained Deep Neural Network for Multiscanner Harmonization.

PhyCHarm was explicitly developed to harmonize multisite and multiscanner MR images. It works to minimize the differences between a T_1_w image obtained from one scanner and a T_1_w image obtained from a different scanner. PhyCHarm works in two steps to generate quantitative maps and harmonized T_1_w images. First, the Quantitative Maps Generator of PhyCHarm generates quantitative T_1_- and M_0_-maps from a T_1_w image obtained on a source scanner. By using the Bloch equation as prior knowledge with the predicted quantitative maps, we can then generate a constrained T_1_w image on a target scanner. Finally, the Harmonization network is used to generate a harmonized T_1_w image.

To evaluate the Quantitative Maps Generator, we compared structural similarity index measure (SSIM) (Wang et al., 2004), peak signal-to-noise ratio (PSNR), and normalized root mean-squared error (NRMSE) techniques. To assess the Harmonization Network, we conducted a comparative analysis with other approaches, including U-Net and Pix2Pix, using metrics such as SSIM and PSNR. We also explored the effects of modifying the weights of the consistency loss in the Quantitative Maps Generator on the ultimate harmonization outcomes in the Harmonization Network. Finally, we compared the gray matter (GM) and white matter (WM) segmentation results generated using each harmonized method through FSL FAST (Smith et al., 2004).

## 2. METHODS

### 2.1 Overall pipeline

The PhyCHarm pipeline is shown in Figure 1. The Quantitative Maps Generator uses the T_1_w image from the source scanner to predict the quantitative T_1_- and M_0_-maps. The constrained T_1_w image is then generated using the estimated T_1_- and M_0_-maps to calculate the T_1_w signal equation for the target scanner, based on the Bloch equation. Finally, the Harmonization Network refines the constrained T_1_w image to closely resemble the T_1_w image from the target scanner. The Quantitative Maps Generator and the Harmonization Network were trained using a 2D U-Net architecture.

**FIGURE 1.**
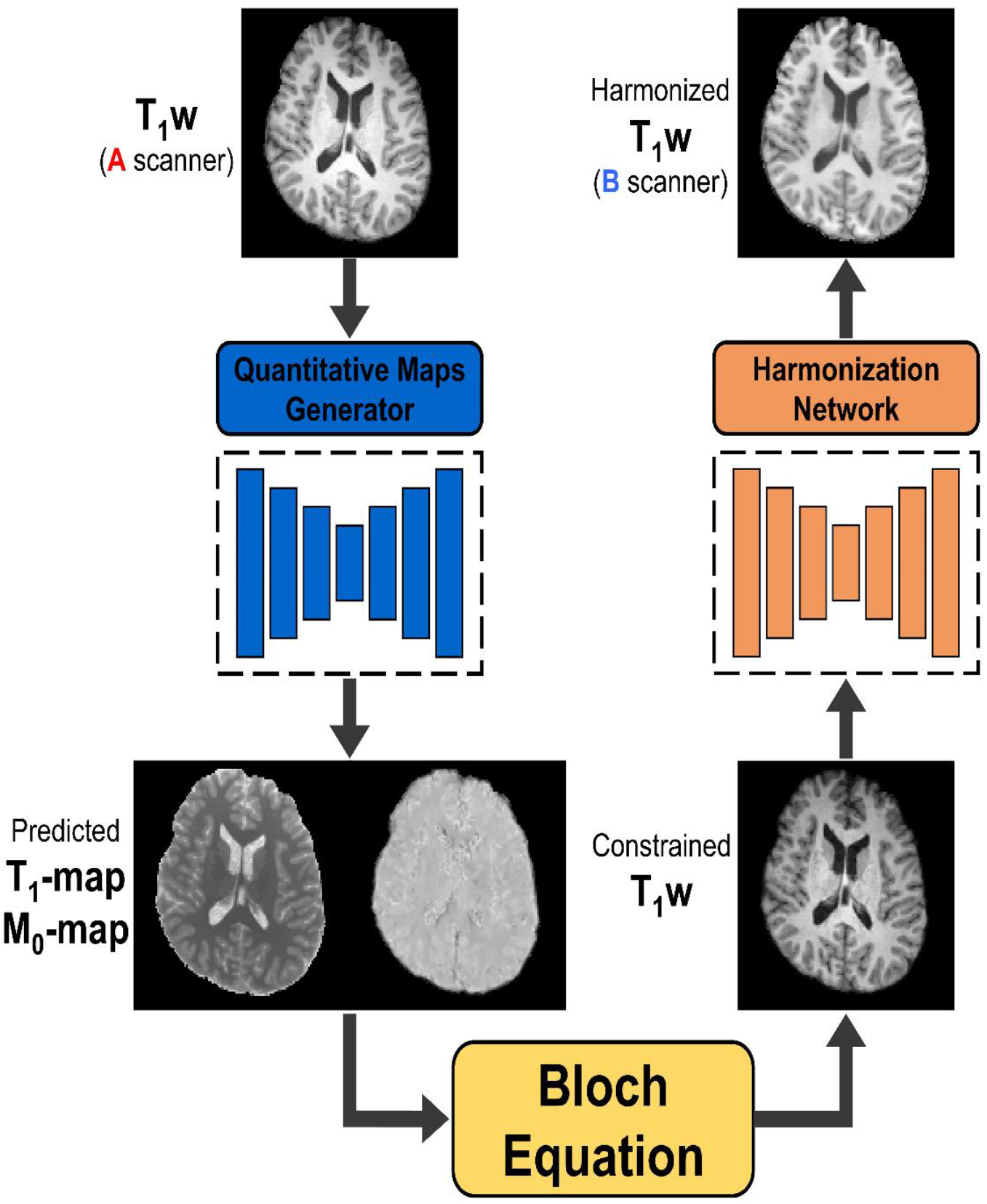
Inference pipeline of PhyCHarm. The T_1_w image acquired from the source scanner (**A** scanner) underwent a series of steps to be transformed into the domain of the T_1_w image acquired from the target scanner (**B** scanner). This process included generating quantitative maps, creating the constrained T_1_w image using the generated quantitative maps and Bloch equation with the target scanner’s scan parameters, and harmonizing the constrained T_1_w image to match the target scanner’s T_1_w image.

### 2.2 Dataset

We used data from a group of 50 healthy individuals included in the MICA-MICs dataset (Royer et al., 2022) to develop the Quantitative Maps Generator. This dataset includes T_1_w images acquired using the MPRAGE sequence (Mugler and Brookeman, 1990), two inversion images (INV1, INV2), T_1_w images, and T_1_-map images obtained from the MP2RAGE sequence (Marques et al., 2010). MPRAGE T_1_w was acquired using a voxel size of 0.8 mm^3^ iso, and an image matrix of 320 × 320 × 224 in the sagittal plane. The repetition time (TR) was 2300 ms; the inversion time (TI) and echo time (TE) were 900 and 2.9 ms, respectively. Imaging data from MP2RAGE were acquired using a voxel size of 0.8 mm^3^ iso, with 320 × 320 × 240 as the sagittal image. The TR was 5000 ms, and the TE was 2.9 ms. The first TI (TI_1_) was 940 ms, and the second TI (TI_2_) was 2830 ms. The flip angles were 4° and 5°. From these datasets, we calculated the M_0_-map using the signal equation of inversion recovery derived from the Bloch equation:

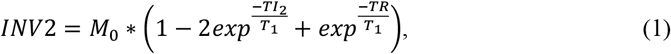

where *M*_0_ is the *M*_0_ map image and *T*_1_ is the *T*_1_ map image acquired from the MP2RAGE sequence.

To train and evaluate the Harmonization Network, we collected traveling T_1_w images. Nine healthy volunteers who provided written consent (IRB approved) were scanned. These images were obtained on three different 3T MR scanners located at various sites: a Trio scanner (Siemens Healthineers, Erlangen, Germany), a SIGNA scanner (GE Medical Systems, Milwaukee, WI, USA), and an Ingenia CS scanner (Philips Healthcare, Best, the Netherlands). The scan parameters for the Harmonization Network dataset are shown in Table S1 (Supplementary Material).

### 2.3. Preprocessing

For the Quantitative Maps Generator dataset, skull stripping was performed using HD-BET (Isensee et al., 2019). To match the image size to 240 × 240 × 224, we cropped the images by applying ANTsPy (Tustison et al., 2021) *crop_image* to the MPRAGE T_1_w, MP2RAGE T_1_-map, and MP2RAGE M_0_-map images.

For the Harmonization Network dataset, the field of view was automatically cropped to remove the lower head and neck using FSL *robustfov*. We then performed skull stripping on the datasets from the three scanners using HD-BET. To achieve a voxel size of 0.8 mm^3^ iso, we used spline interpolation to perform resampling on the GE dataset. The image size of the resampled GE dataset was then modified to a matrix size of 240 × 240 × 224 by cropping and padding using ANTsPy. To achieve proper alignment, we used affine registration techniques to accurately map the spatial orientation of the Siemens and Philips datasets to correspond to that of the GE dataset by using ANTsPy. All vendor datasets underwent N4 bias correction to correct for B_0_ inhomogeneities.

### 2.4. Training networks

Figure 2 shows the training pipeline of the Quantitative Maps Generator and the Harmonization Network. A total of 4,480 slices from 20 individuals were initially used in the training process for the Quantitative Maps Generator; 1,120 slices from five individuals were subsequently incorporated for validation. We used the ADAM optimizer (Kingma and Ba, 2015). The total epoch was set to 500. The Quantitative Maps Generator was trained with the total loss (*L*_*total*_), which is a calculated combination of the reconstruction loss (*L*_*T*1_, *L*_*M*0_) and the consistency loss (*L*_*cons*_). *L*_*T*1_, *L*_*M*0_, *L*_*cons*_ were updated based on the mean squared error (MSE) loss:

**FIGURE 2.**
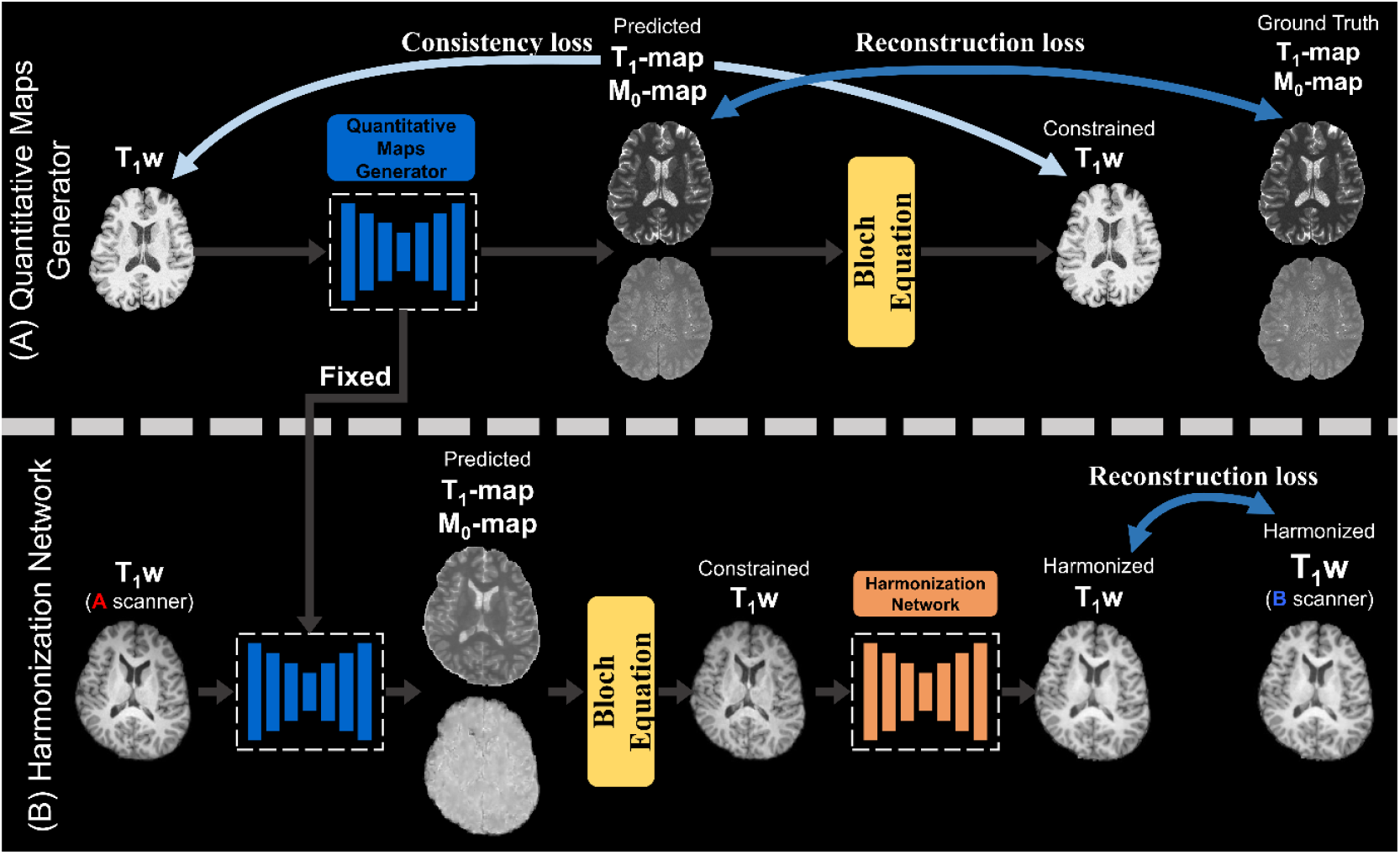
Training pipeline. PhyCHarm involves two deep neural networks: (A) the Quantitative Maps Generator and (B) the Harmonization Network. The Quantitative Maps Generator was trained to generate T_1_- and M_0_-maps. To train the Harmonization Network, the pretrained Quantitative Maps Generator was used to generate the 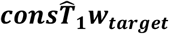, which is the input of the Harmonization Network. The Bloch equation was used in two ways: (1) to generate ***consT***_***1***_***W*** to calculate the additional loss (consistency loss) for the Quantitative Maps Generator and (2) to generate the input for the Harmonization Network (the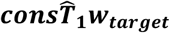).

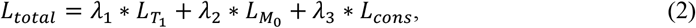

where *λ*_1_= 1, *λ*_2_ = 1, and *λ*_3_ = _1_*e*^−6^. The weight values of all losses were determined by comparing four cases, defined as follows: Case 1, *λ*_1_ : *λ*_2_ : *λ*_3_ = 1 : 1: 0 (without Bloch-equation constraints); Case 2, *λ*_1_ : *λ*_2_ : *λ*_3_ = 0.5 : 0.5 : _1_*e*^−6^; Case 3, *λ*_1_ : *λ*_2_ : *λ*_3_ = 1 : 1 : _1_*e*^−8^; and Case 4, *λ*_1_ : *λ*_2_ : *λ*_3_ = 1 : 1 : _1_*e*^−6^, where *λ*_1_ represents the weight assigned to the T_1_-map loss, *λ*_2_ represents the M_0_-map loss, and *λ*_3_ represents the consistency loss.

The reconstruction loss was determined by comparing the predicted T_1_- and M_0_-maps with the true T_1_- and M_0_-maps. Calculating the consistency loss involved comparing the input T_1_w image with the constrained T_1_w image. The constrained T_1_w image was generated using Eqs. 3 through 6 as part of the training process.

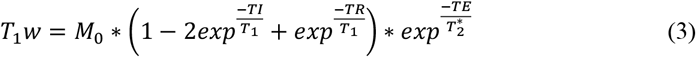

In Eq. 3, the T_1_w equation represents the inversion-recovery signal equation in a general manner.

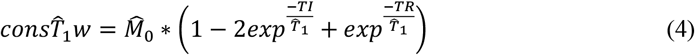

In Eq. 4, 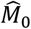 is a predicted M_0_-map, 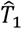 is a predicted T_1_-map, and *cons* 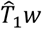 is the constrained T_1_w image that was calculated using 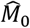 and 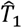 by ignoring 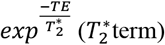. In this equation, TI = 1000 ms and TR = 2300 ms.

We obtained a corrective term using the relationship between the input T_1_w and 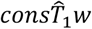 as shown in Eq. 5:

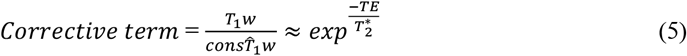

The corrective term included 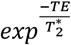. A recalculation of the constrained T_1_w (*consT*_1_*w*) was then performed by incorporating the corrective term, 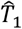, and 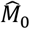, as shown in Eq. 6:

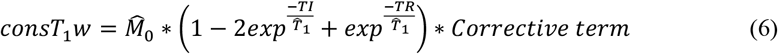

When training the Harmonization Network, 672 slices from three individuals were initially used in the training process for the Quantitative Maps Generator, and 224 slices from one individual were subsequently incorporated for validation. For inference, we used 1,120 slices from five individuals. The pretrained Quantitative Maps Generator was used to generate *T*_1_- and *M*_0_-maps as all of its parameters were fixed. Given the T_1_w image of the source scanner (input domain, T_1_w_input_), the Quantitative Maps Generator predicted the T_1_- and M_0_-maps corresponding to the T_1_w_input_. The constrained 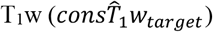 was then generated using the scan parameters of the target scanner. The Harmonization Network was trained to generate a harmonized T_1_w image from this constrained T_1_w image. To acquire 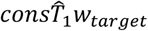 using the corrective term, we first used the scan parameters of the source scanner to calculate the constrained T_1_w of the source scanner 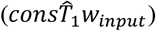:

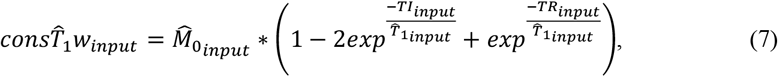

where TI_*input*_ and TR_*input*_ are the TI and TR of the source scanner and 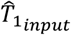 and 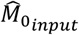 are the predicted T_1_- and M_0_-maps from the Quantitative Maps Generator when the input data is the T_1_w of the source scanner (*T*_1_*w*_*input*_). We then obtained a corrective term:

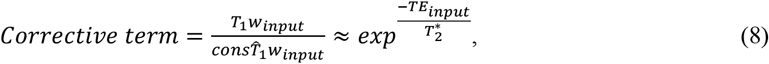

which included the 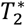-related term 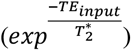. A pseudo 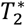 map 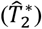 was then calculated:

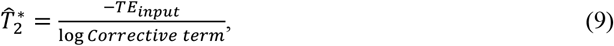

Where

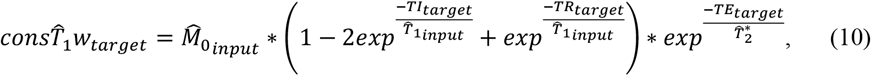

which includes the scan parameters of the target scanner (TI_*target*_, TR_*target*_, and TE_*target*_), defined as the TI, TR, and TE of T_1_w acquired from the target scanner.

We trained the Harmonization Network for three harmonization pairs. These pairs included GE and Siemens, Siemens and Philips, and Philips and GE. To mitigate the issue of overfitting caused by the small number of subjects, we applied a four-fold cross-validation approach and dropout with a probability of 0.1. Harmonization Network training was updated by incorporating reconstruction loss using the MSE loss function. With this approach, we aimed to minimize the disparity between the harmonized T_1_w image and the ground truth T_1_w image. We employed the ADAM optimizer with a learning rate of 0.001, and we established a total of 200 training epochs.

### 2.5. Network architecture

Implementation of the Quantitative Maps Generator and Harmonization Network was performed using 2D U-Net. The 2D U-Net encoder was implemented using output channels of varying sizes: 16, 32, 64, 128, and 256. The 2D U-Net encoder was constructed with interconnected blocks, using max-pooling layers with a kernel size and stride of 2. Each block was composed of two pairs of Conv2d-LeakyRELU, and instance normalization was incorporated between Conv2d and LeakyReLU. The 2D U-Net decoder used input channel numbers 128, 64, 32, and 16. The decoder also used Conv2d-LeakyRELU blocks with up-sampling, with a kernel size and stride of 2.

### 2.6. Evaluation

For the Quantitative Maps Generator, quantitative evaluation scores (SSIM, PSNR, and NRMSE) were analyzed for 5,600 slices from 25 participants from the open dataset. We compared the evaluation scores for Case 1, Case 2, Case 3, and Case 4.

For the Harmonization Network, five traveling subject datasets were additionally acquired for an external inference dataset. A total of 1,120 slices from five traveling subjects were used for inference. PhyCHarm results were compared with results from unconstrained U-Net, Pix2Pix, and CALAMITI using SSIM and PSNR. Three harmonization pairs were again examined: GE and Siemens, Siemens and Philips, and Philips and GE. Histogram analysis was also conducted.

For the downstream task, volumetric assessments were performed on the segmentation masks for GM and WM, which were generated using FSL FAST. We compared the Dice scores for GM and WM segmentation masks between the ground truth and the harmonized results. We additionally compared the volumetric discrepancies in GM and WM for each scanner pair before and after harmonization.

### 2.7. Statistical analysis

All statistical evaluations were performed using Python (Version 3.8.10; Python Software Foundation) and the SciPy library. We conducted the Wilcoxon signed-rank test on the Dice scores, using a significance level of *p* < 0.05.

## 3. RESULTS

### 3.1. Quantitative maps generator

The Quantitative Maps Generator was able to generate T_1_- and M_0_-maps with comparable quality to the ground truth, as highlighted in the difference maps shown in Figure 3.

**FIGURE 3.**
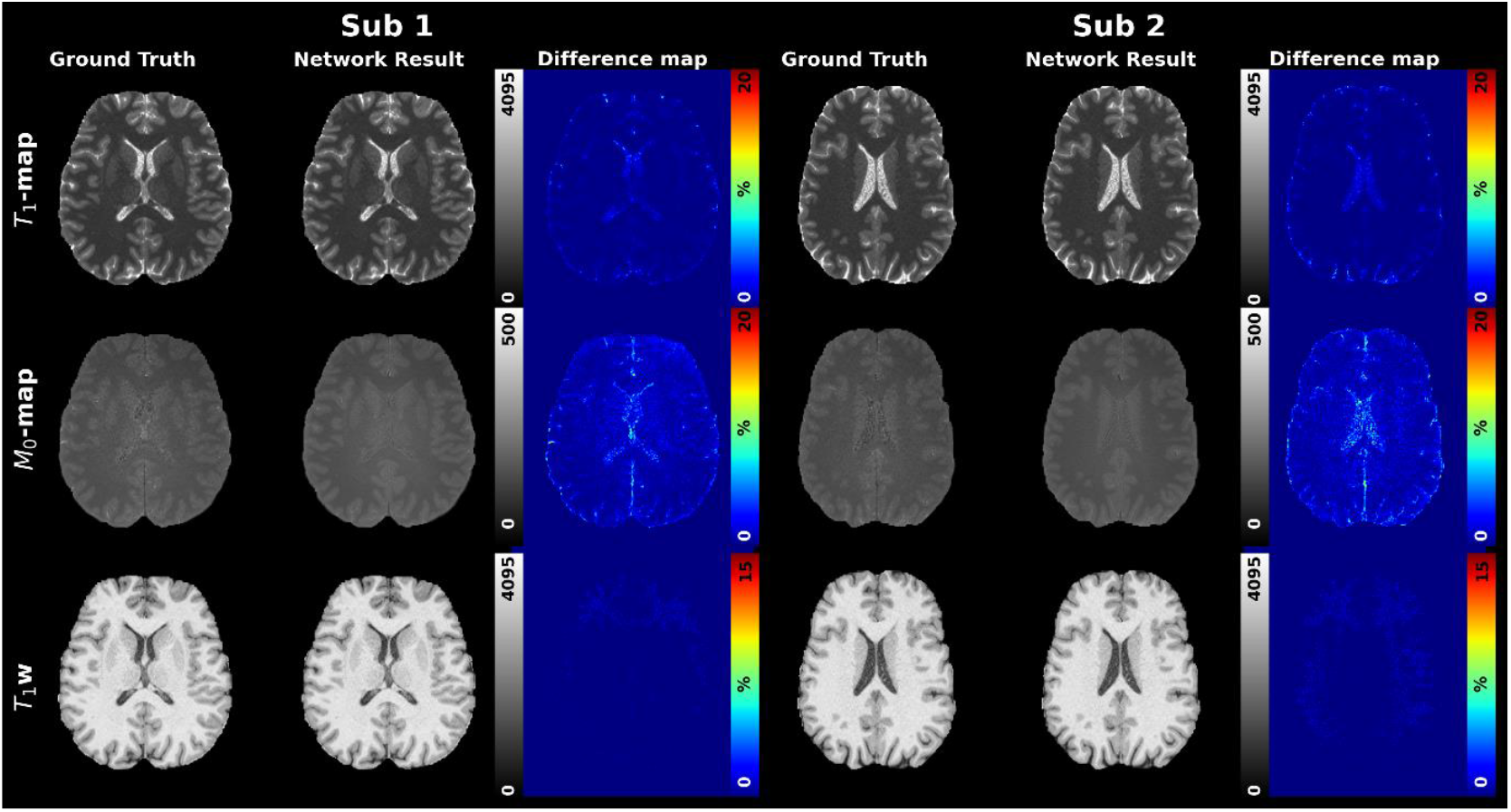
Inference results for the Quantitative Maps Generator. The results for two representative subjects are displayed. The ground truths for the T_1_-map, M_0_-map, and T_1_w images are shown in the Ground Truth column. The Network Result column shows the generated T_1_-map, M_0_-map, and the constrained T_1_w image. The Difference map column shows the difference between the ground truth and the inference results for the T_1_-and M_0_-map images, as well as the difference between the ground truth T_1_w image and the constrained T_1_w image.

Table 1 presents the quantitative evaluation results for the Quantitative Maps Generator for each of the four weight value cases defined earlier. Initially, a comparison was made between Cases 2 and 4 to ascertain the most optimal weight values for the loss of the T_1_-(*λ*_1_) and M_0_-maps (*λ*_2_). It was noted that when *λ*_1_ and *λ*_2_ were set to 0.5 (Case 2), the SSIM and PSNR were lower and the NRMSE was higher than when *λ*_1_ and *λ*_2_ were set to 1 (Case 4). Subsequently, after setting the weight values for these losses as 1, we evaluated the three distinct weight values for the consistency loss (*λ*_3_): 0 (Case 1), _1_*e*^−8^ (Case 3), and _1_*e*^−6^ (Case 4). Case 4 exhibited the highest SSIM and PSNR values while demonstrating the lowest NRMSE value. The SSIM and PSNR values for the T_1_-map were highest in Case 4, whereas the values for the M_0_-map were highest in Case 3. Because the evaluation scores between Cases 3 and 4 showed a larger difference for the T_1_-map than for the M_0_-map, we used the Quantitative Maps Generator trained with the weight values from Case 4 to train the Harmonization Network.

**TABLE 1.** Quantitative evaluation scores for the Quantitative Maps Generator (inference).

|  |  | Case 1 | Case 2 | Case 3 | Case 4 |
| --- | --- | --- | --- | --- | --- |
| T<sub>1</sub>-map | SSIM | 0.9980 | 0.9796 | 0.9980 | **0.9996** |
|  | PSNR | 42.3358 | 29.0202 | 41.6810 | **50.2640** |
|  | NRMSE | 0.0565 | 0.2651 | 0.0592 | **0.0240** |
| M<sub>0</sub>-map | SSIM | 0.9457 | 0.9398 | **0.9462** | 0.9459 |
|  | PSNR | 31.4074 | 27.4536 | **32.7790** | 30.6528 |
|  | NRMSE | 0.1223 | 0.2058 | **0.1113** | 0.1154 |

*Note:* The bold numbers represent the highest SSIM and PSNR values and the lowest NRMSE value. SSIM = structural similarity index measure; PSNR = peak-signal-to-noise ratio; NRMSE = normalized root mean-squared error.

### 3.2. Harmonization network

Figure 4 shows the qualitative comparison of validation results for PhyCHarm versus the results for U-Net, Pix2Pix, and CALAMITI. U-Net and Pix2Pix exhibited varying performance depending on the input domain. CALAMITI demonstrated reduced scanner bias compared to U-Net and PhyCHarm across all input and target domains. Additionally, CALAMITI generated specific features that appeared less visually smoothed, especially within the WM. PhyCHarm demonstrated consistent accuracy in harmonization results across all input domains.

**FIGURE 4.**
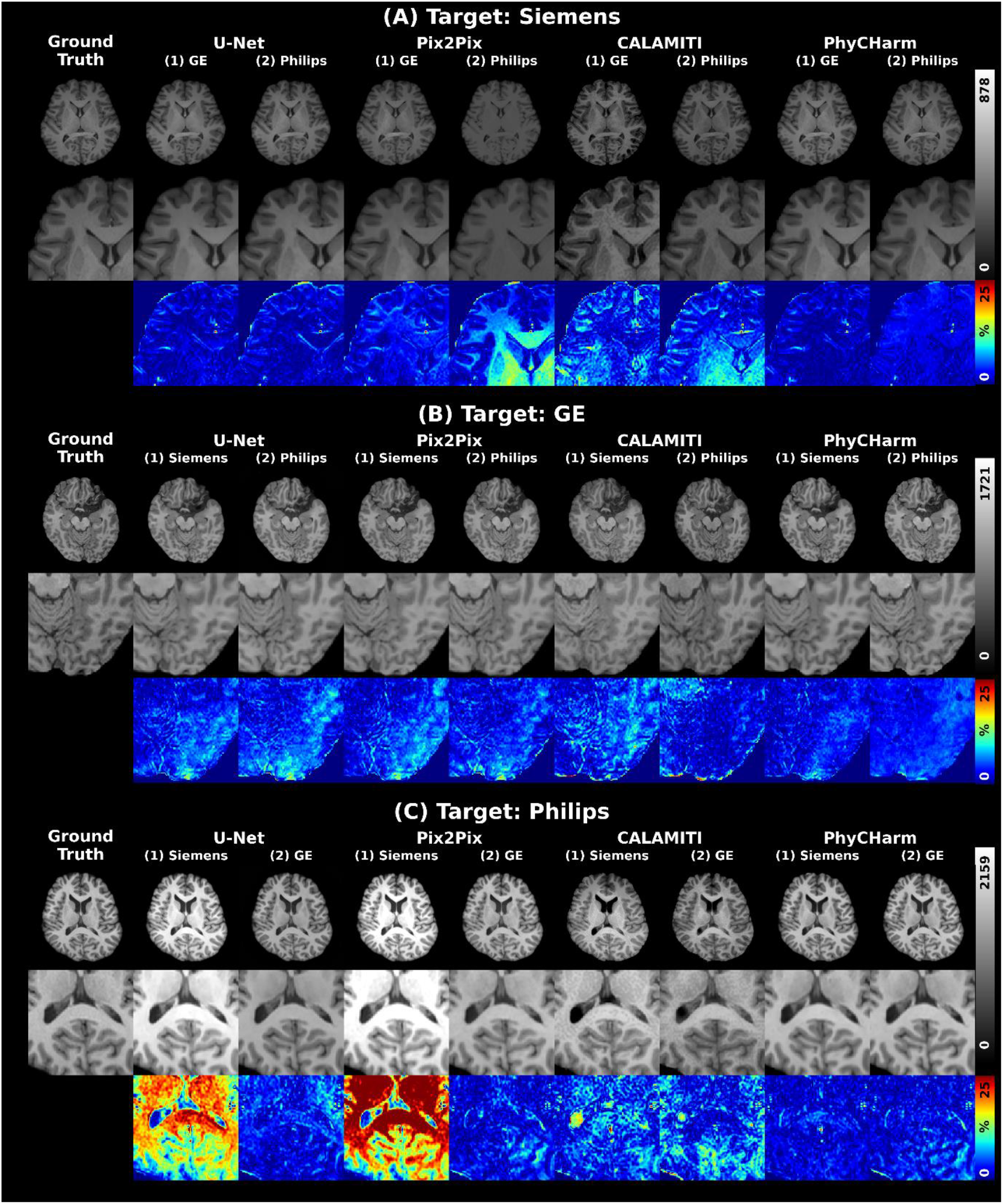
Visual comparison of harmonization results for the validation set. The first rows in (A), (B), and (C) show each ground truth and the harmonization results from U-Net, Pix2Pix, CALAMITI, and PhyCHarm. The second rows in (A), (B), and (C) display the zoomed-in results of the first rows. The third rows in (A), (B), and (C) show the error maps. (A) Results achieved for the Siemens target domain with the (1) GE input domain and (2) Philips input domain. U-Net shows comparable harmonization quality when the target domain is Siemens. Pix2Pix demonstrates varying accuracy depending on the input domain. CALAMITI shows consistent quality regardless of the input domain. PhyCHarm maintains stable accuracy across both input domains. (B) Results achieved for the GE target domain with the (1) Siemens input domain and (2) Philips input domain. When comparing the cropped area for (1), U-Net and Pix2Pix tend to generate visually smoother images than CALAMITI and PhyCHarm. PhyCHarm shows the lowest error rate. (C) Results achieved for the Philips target domain with the (1) Siemens input domain and (2) GE input domain. In these harmonization cases, U-Net and Pix2Pix show inconsistent accuracy depending on the input domains. CALAMITI shows consistent quality regardless of the input domain. PhyCHarm demonstrates the lowest error rate.

Figure 5 shows the qualitative comparison of inference results using the external inference dataset. U-Net showed consistent accuracy only when the target domain was GE. When the target domain was Siemens and Philips, U-Net and Pix2Pix exhibited different intensity values compared to the ground truth and blurred structural features. CALAMITI demonstrated better consistency in performance than U-Net and Pix2Pix. CALAMITI also generated specific features that highlighted fine structures within the WM across all input domains. PhyCHarm demonstrated the lowest difference from the ground truth across all input domains.

**FIGURE 5.**
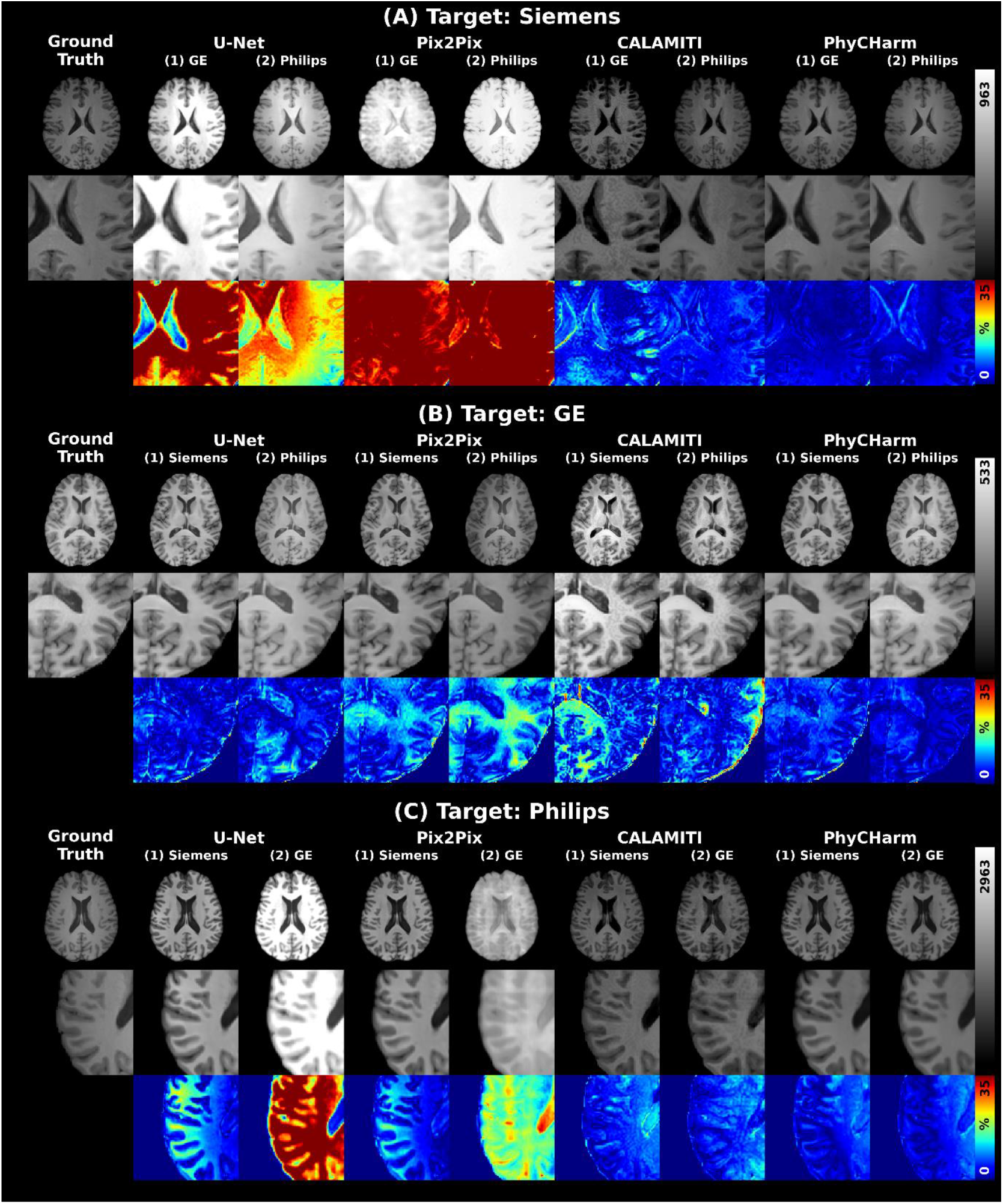
Visual comparison of harmonization results for the external inference set. The first rows in (A), (B), and (C) show each ground truth and the harmonization results from U-Net, Pix2Pix, CALAMITI, and PhyCHarm. The second rows in (A), (B), and (C) display the zoomed-in results of the first rows. The third rows in (A), (B), and (C) show the error maps. (A) Results achieved for the Siemens target domain with the (1) GE input domain and (2) Philips input domain. U-Net and Pix2Pix demonstrate varying accuracy depending on the input domain, whereas CALAMITI and PhyCHarm maintain stable accuracy across both input domains. PhyCHarm demonstrates the lowest error rate for the external inference dataset. (B) Results achieved for the GE target domain with the (1) Siemens input domain and (2) Philips input domain. U-Net demonstrates a lower error rate than Pix2Pix and CALAMITI. U-Net and Pix2Pix tend to generate smoother images in the Philips to GE harmonization case. CALAMITI shows less smoothed features than U-Net and Pix2Pix, but the contrast of CALAMITI results is slightly different from the ground truth. PhyCHarm demonstrates stability, showing consistent accuracy regardless of the input domain. (C) Results achieved for the Philips target domain with the (1) Siemens input domain and (2) GE input domain. U-Net and Pix2Pix show the highest error rate. CALAMITI shows similar contrast and a lower error rate when compared with U-Net and Pix2Pix. However, a darker cerebrospinal fluid intensity is observed with CALAMITI than with the ground truth. PhyCHarm shows the lowest error rate.

In quantitative evaluations, U-Net, Pix2Pix, CALAMITI, and PhyCHarm showed improved SSIM and PSNR values when compared to the method without harmonization in the validation dataset (Table S2 in Supplementary Material). PhyCHarm reveals improved evaluation scores compared to the method without harmonization, U-Net, Pix2Pix, and CALAMITI. For the external inference dataset (Table 2), the performance of U-Net and Pix2Pix varied depending on the input domains, whereas CALAMITI and PhyCHarm both demonstrated consistent evaluation scores. In addition to the quantitative evaluation, we conducted a histogram analysis for the external inference dataset, as shown in Figure S1 (Supplementary Material). PhyCHarm showed the histogram most comparable to that obtained from the ground truth of the target domain.

**TABLE 2.** Quantitative evaluation scores for harmonization methods (inference).

| Harmonization method | GE → Siemens | Philips → Siemens | Siemens → GE | Philips → GE | Siemens → Philips | GE → Philips |
| --- | --- | --- | --- | --- | --- | --- |
|  | SSIM |  |  |  |  |  |
| Without harmonization | 0.8791 | 0.9250 | 0.8806 | 0.9092 | 0.9250 | 0.9076 |
| U-Net | 0.8357 | 0.9237 | **0.9474** | 0.9447 | 0.9282 | 0.8712 |
| Pix2Pix | 0.8380 | 0.8737 | 0.9423 | 0.9456 | 0.9286 | 0.8725 |
| CALAMITI | 0.9055 | 0.9164 | 0.9337 | 0.9401 | 0.9186 | 0.9236 |
| PhyCHarm | **0.9579** | **0.9434** | 0.9468 | **0.9772** | **0.9362** | **0.9362** |
|  | PSNR |  |  |  |  |  |
| Without harmonization | 18.585 | 26.0914 | 19.3282 | 21.4230 | 26.0914 | 20.7115 |
| U-Net | 16.1404 | 26.5954 | 27.3701 | 23.5538 | 26.6575 | 15.0503 |
| Pix2Pix | 14.0342 | 20.3272 | 25.5793 | 25.1624 | 28.2067 | 17.2583 |
| CALAMITI | 24.4774 | 26.6101 | **28.3925** | 29.0506 | 25.7760 | **27.0543** |
| PhyCHarm | **29.9030** | **29.9601** | 25.2000 | **29.3897** | **29.6438** | 22.7095 |

*Note:* → denotes harmonizing the input domain to the target domain. The bold numbers represent the highest SSIM and PSNR values. SSIM and PSNR for the method without harmonization were calculated between the original data from the target domain and the original data from the input domain. SSIM and PSNR for the method with harmonization were measured between the original data from the target domain and the harmonized data from the input domain. SSIM = structural similarity index measure; PSNR = peak-signal-to-noise ratio

Figure S2 (Supplementary Material) provides a visual representation of the progression of training loss during Harmonization Network training, focusing specifically on the impact of weight values on the consistency loss (*λ*_3_) from the Quantitative Maps Generator. There was a significant decrease at the 25th epoch when *λ*_3_ was _1_*e*^−6^ (Case 4) compared to when *λ*_3_ was 0 (Case 1). Throughout the training of the Harmonization Network for GE to Siemens harmonization, there was an immediate noticeable reduction in loss. The harmonization from Philips to GE demonstrated the impact of the consistency of the Bloch equation on early loss reduction. This effect was particularly evident around the 30th epoch.

### 3.3. GM and WM segmentation

Table S3 (Supplementary Material) and Table 3 show the mean Dice scores for GM and WM segmentation results obtained using FSL FAST, based on the validation dataset and the external inference dataset, respectively. In the segmentation results using the validation dataset, PhyCHarm achieved the highest Dice scores for both GM and WM across all harmonization cases. For the segmentation results using the external inference dataset, CALAMITI showed the highest Dice scores for WM segmentation in one harmonization case (Siemens to GE), whereas PhyCHarm achieved the highest Dice scores for GM and WM segmentation in three harmonization cases (GE to Siemens, Philips to GE, and GE to Philips). Figures 6 and 7 show the qualitative comparison of FSL FAST GM and WM segmentation results using the external inference dataset. PhyCHarm demonstrated minimal disparities in the absolute difference maps.

**TABLE 3.** Dice scores (inference)

| Harmonization method | GE → Siemens | Philips → Siemens | Siemens → GE | Philips → GE | Siemens → Philips | GE → Philips |
| --- | --- | --- | --- | --- | --- | --- |
|  | **GM** |  |  |  |  |  |
| Without harmonization | 0.8310 | **0.8330** | **0.8268** | 0.8061 | **0.8330** | 0.8061 |
| U-Net | 0.6060<br>($p < 0.05$) | 0.7107<br>($p < 0.05$) | 0.7916<br>($p < 0.05$) | 0.7304<br>($p < 0.05$) | 0.7630<br>($p < 0.05$) | 0.7208<br>($p < 0.05$) |
| Pix2Pix | 0.6580<br>($p < 0.05$) | 0.6278<br>($p < 0.05$) | 0.7752<br>($p < 0.05$) | 0.7084<br>($p < 0.05$) | 0.7591<br>($p < 0.05$) | 0.7028<br>($p < 0.05$) |
| CALAMITI | 0.7336<br>($p < 0.05$) | 0.7526<br>($p < 0.05$) | 0.7071<br>($p < 0.05$) | 0.7123<br>($p < 0.05$) | 0.7367<br>($p < 0.05$) | 0.7249<br>($p < 0.05$) |
| PhyCHarm | **0.8377\***<br>($p < 0.05$) | 0.7958<br>($p < 0.05$) | 0.7846<br>($p < 0.05$) | **0.8555\***<br>($p < 0.05$) | 0.8034<br>($p < 0.05$) | **0.8313\***<br>($p < 0.05$) |
|  | **WM** |  |  |  |  |  |
| Without harmonization | 0.8980 | **0.8380** | 0.8307 | 0.8478 | **0.8858** | 0.8478 |
| U-Net | 0.7480<br>($p < 0.05$) | 0.7738<br>($p < 0.05$) | 0.8266<br>($p < 0.05$) | 0.8172<br>($p < 0.05$) | 0.8830<br>($p < 0.05$) | 0.6259<br>($p < 0.05$) |
| Pix2Pix | 0.8347<br>($p < 0.05$) | 0.7631<br>($p < 0.05$) | 0.8135<br>($p < 0.05$) | 0.8205<br>($p < 0.05$) | 0.8738<br>($p < 0.05$) | 0.7060<br>($p < 0.05$) |
| CALAMITI | 0.8587<br>($p < 0.05$) | 0.7536<br>($p < 0.05$) | **0.8501\***<br>($p < 0.05$) | 0.8320<br>($p < 0.05$) | 0.8262<br>($p < 0.05$) | 0.8520*<br>($p < 0.05$) |
| PhyCHarm | **0.8988\***<br>($p < 0.05$) | 0.7963<br>($p < 0.05$) | 0.8294<br>($p < 0.05$) | **0.9039\***<br>($p < 0.05$) | 0.8669<br>($p < 0.05$) | **0.8934\***<br>($p < 0.05$) |

*Note:* Dice scores are displayed as mean values. Dice scores for the method without harmonization were calculated between the original data from the target domain and the original data from the input domain. Dice scores for the method with harmonization were calculated between the original data from the target domain and the harmonized data from the input domain. A Wilcoxon signed-rank test was performed to compare the Dice scores for the method without harmonization with those for the other harmonization methods. Scores marked with \* indicate significantly higher Dice scores compared to the method without harmonization ( $p < 0.05$ ). The bold values denote the highest Dice scores. GM = gray matter; WM = white matter.

**FIGURE 6.**
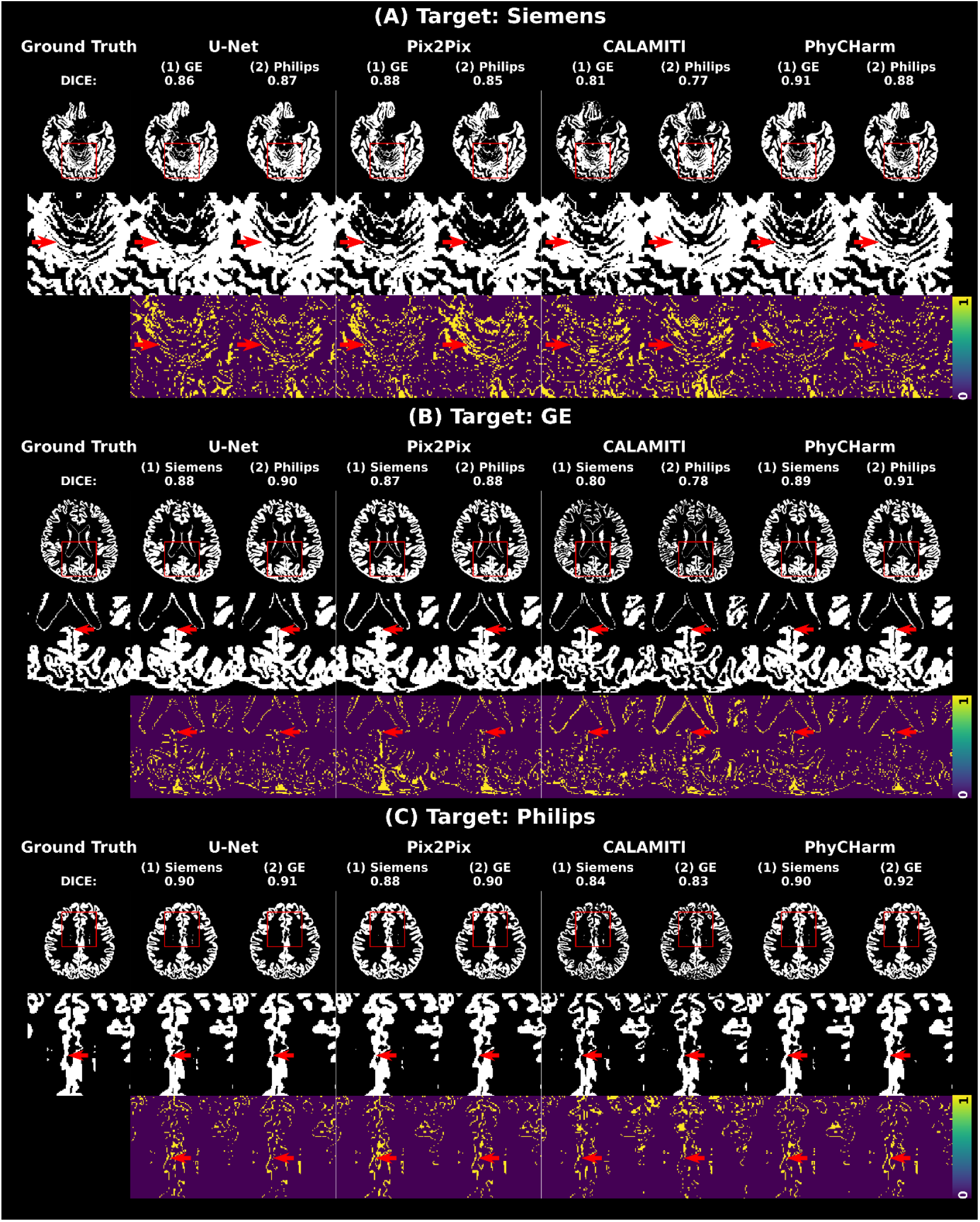
Results of segmentation for gray matter (GM). The first rows in (A), (B), and (C) show the FSL FAST segmentation results for GM from the ground truth and the harmonization results from U-Net, Pix2Pix, CALAMITI, and PhyCHarm. The source scanner name and Dice score are indicated above the brain mask images in the first rows. The second rows in (A), (B), and (C) display the zoomed-in results of the first rows, and the third rows show the zoomed-in difference maps. The harmonization results with PhyCHarm are comparable to the ground truth, particularly in the areas highlighted by arrows.

**FIGURE 7.**
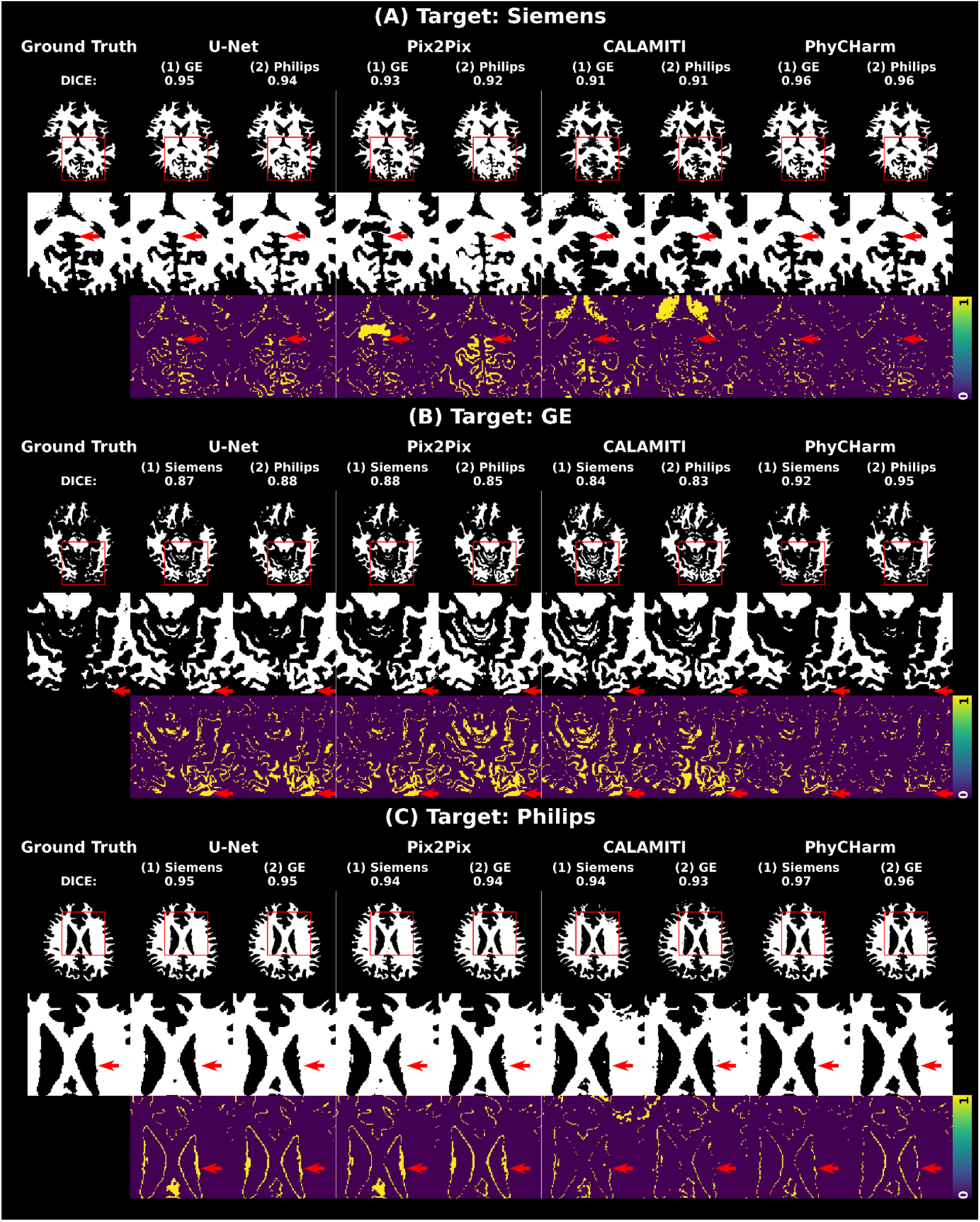
Results of segmentation for white matter (WM). The first rows in (A), (B), and (C) show the FSL FAST segmentation results for WM from the ground truth and the harmonization results from U-Net, Pix2Pix, CALAMITI, and PhyCHarm. The source scanner name and Dice score are indicated above the brain mask images in the first rows. The second rows in (A), (B), and (C) display the zoomed-in results of the first rows, and the third rows show the zoomed-in difference maps. PhyCHarm demonstrates the highest Dice score, particularly in the areas highlighted by arrows.

Figure S3 (Supplementary Material) and Figure 8 show the GM and WM volume differences between the input and target domains for the validation dataset and the external inference dataset, respectively. The volume was measured from the FSL FAST segmentation masks. In the validation results, PhyCHarm notably reduced the GM volumetric difference in the Siemens to GE, Philips to GE, Siemens to Philips, and GE to Philips harmonization cases. In the inference results using the external dataset, U-Net showed an increased volumetric difference in all harmonization cases, contrary to the trends observed in the validation results. Pix2Pix reduced the WM volumetric difference in the Philips to GE harmonization case. CALAMITI reduced the WM volumetric difference in the GE to Siemens and the Philips to Siemens harmonization cases. PhyCHarm reduced the WM volumetric difference in all harmonization cases except for the Siemens to GE harmonization case.

**FIGURE 8.**
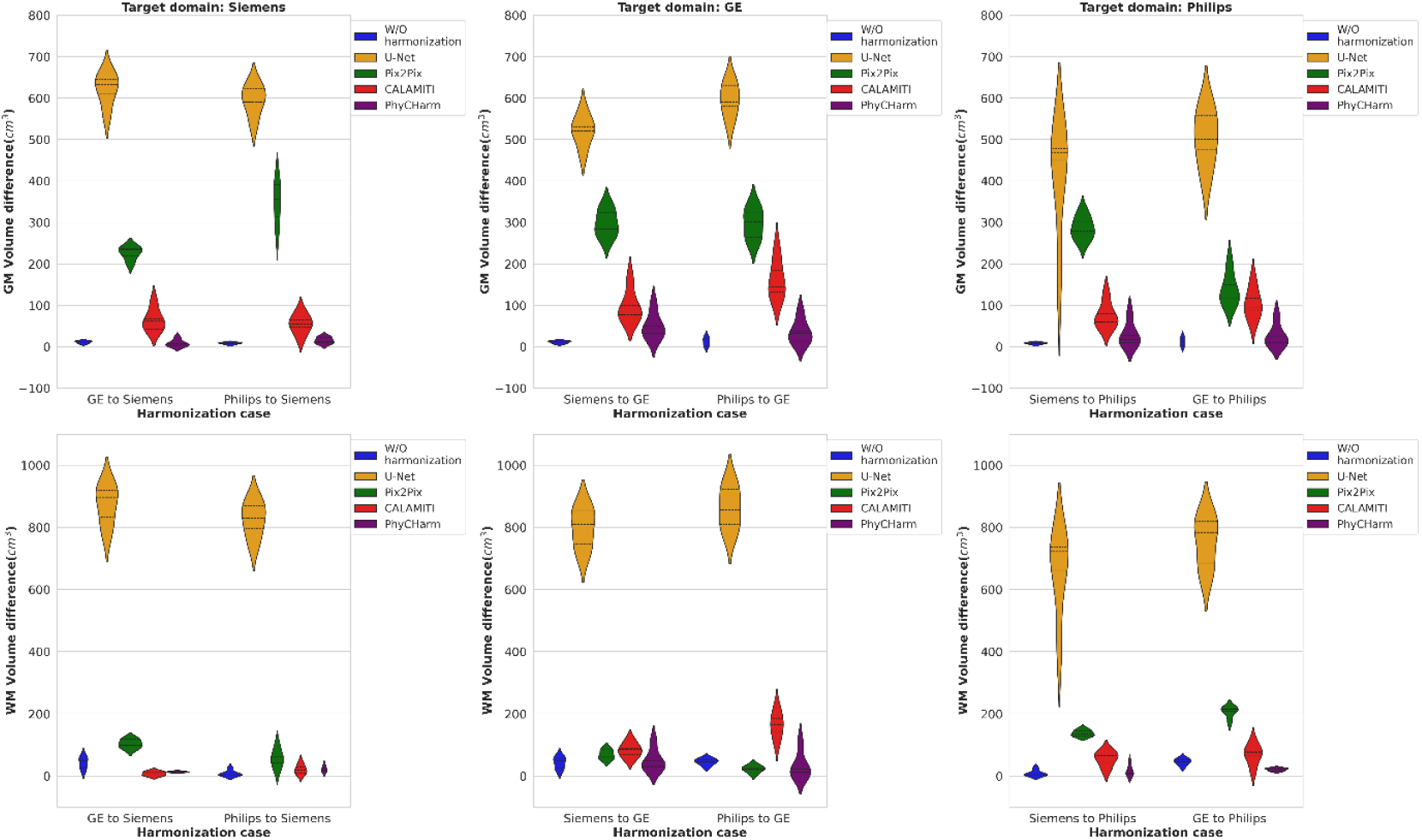
Gray matter (GM) and white matter (WM) volumetric differences in the inference results using the external inference dataset. The first row shows the GM volumetric differences; the second row shows the WM volumetric differences. The volumetric difference was calculated between the input domain and target domain volumes. “W/O harmonization” represents the results obtained without applying harmonization. U-Net shows increased GM and WM volumetric differences across all harmonization cases. Pix2Pix shows increased GM volumetric differences across all harmonization cases, whereas the WM volumetric difference is decreased in the Philips to GE harmonization case. CALAMITI shows increased GM volumetric differences in all harmonization cases, whereas the WM volumetric difference is reduced in the GE to Siemens harmonization case. PhyCHarm shows reduced GM volumetric difference in the GE to Siemens harmonization case and reduced WM volumetric differences in all harmonization cases with the exception of GE to Siemens.

## 4. DISCUSSION

Theoretically, it is possible to synthesize all desired contrasts from T_1_-, T_2_-, and M_0_-maps using the Bloch equation. However, there are practical constraints in synthesizing images using this method. Multiple datasets and scans are required to calculate quantitative maps, such as multi-inversion time datasets for T_1_ mapping and multiecho datasets for T_2_ mapping. This lengthens the total acquisition time and limits the clinical use of this technique.

To overcome these constraints, we have developed the PhyCHarm method, which employs a deep neural network and the Bloch equation to harmonize MR images. The Bloch equation is used as the training constraint for the Quantitative Maps Generator, which provides prior knowledge to the deep neural networks related to the correlation between the input T_1_w image and ground truth data of the T_1_- and M_0_-maps during training. This Quantitative Maps Generator can generate T_1_- and M_0_-maps based on an acquired T_1_w image without the need for additional data acquisition. In this study, the Quantitative Maps Generator required 20 seconds to generate the T_1_- and M_0_-maps for one individual. This speed was measured on a platform consisting of a single RTX 3090 GPU (GPU-RAM capacity of 24 GB) and eight workers for the data loader.

To train the Harmonization Network, we used the Bloch equation to minimize the feature difference due to scan parameters between input and target scans. With this method, we observed a rapid reduction of loss when the weight value of the consistency loss was _1_*e*^−6^. This rapid reduction of loss resulted in a lower loss value at the 200th epoch than when the weight value of the consistency loss was 0. This finding suggests that physics constraints help prevent local minima by reducing the factors that influence the scanner effect. With this technique, the controllable factors contributing to the scanner effect are minimized.

To compare PhyCHarm with state-of-the-art methods, we used CALAMITI in a supervised manner, as applied in MISPEL.^33^ MISPEL is also considered a state-of-the-art method; however, it works by reducing the disparity between the input and target domains rather than by employing a direct mapping (commonly referred to as domain shift) from the input to the target domain, which was the focus of our study. We found that when compared with U-Net and Pix2Pix, CALAMITI demonstrated improved consistency in both qualitative comparisons and evaluation scores when using the external inference dataset. In contrast, U-Net and Pix2Pix demonstrated notable inconsistencies, particularly with the external inference dataset, likely due to overfitting to the validation dataset. Although CALAMITI showed consistent performance for the external inference dataset in this study, it also generated specific image features that led to less smoothed images than were seen with PhyCHarm. This finding has not been reported in previous studies, perhaps in part because we used a smaller number of datasets. Despite the generation of these features, CALAMITI demonstrated improved SSIM and PSNR compared to U-Net and Pix2Pix and outperformed PhyCHarm in the PSNR comparison for some harmonization cases.

PhyCHarm demonstrated better segmentation consistency and accuracy when compared with U-Net, Pix2Pix, and CALAMITI. When applied to the external inference dataset, PhyCHarm improved segmentation accuracy in 3 out of 6 cases. This indicates that PhyCHarm has the advantage of enhancing harmonization performance while maintaining the structural characteristics of GM and WM. This suggests that PhyCHarm could serve as an effective preprocessing step for anatomical analysis of healthy brains, as PhyCHarm reduces disparities caused by differences in scanners and sites. Applying physics constraints allows deep neural networks to undergo training that preserves primary anatomical features and intricate characteristics that may be overlooked during resampling within deep neural network architectures. Future work could further improve the consistency and accuracy for the external inference dataset by integrating additional constraint components such as GM or WM masks.

This study had several limitations. First, the Quantitative Maps Generator and the Harmonization Network were trained using a 2D architecture because of limited GPU RAM; this prevented the implementation of a 3D network. If the two networks and additional deep neural networks for segmentation or other downstream tasks are combined, a 2.5D or 3D network may be needed for accurate segmentation accuracy. Second, the datasets used for both networks did not include patient data. Because the T_1_ and M_0_ values for lesions differ from the values seen in healthy controls, newly fine-tuned parameters for both networks may be required for patient datasets. Third, the dataset used for the Harmonization Network lacks diversity in age and racial representation. Applying our framework to different age groups or races would necessitate an optimal fine-tuning approach. The use of foundation models may improve the generalizability of the Harmonization Network, as foundation models for the medical domain have demonstrated enhanced generalizability with multimodalities and small-scale medical datasets (Azizi et al., 2023). In future work, we plan to investigate the methodology of combining the PhyCHarm framework with foundation models.

## 5. CONCLUSION

In this study, we proposed a physics-constrained deep neural network for MR harmonization, PhyCHarm, to generate T_1_- and M_0_-maps in the Quantitative Maps Generator and to harmonize brain T_1_w images acquired from different scanners or sites in the Harmonization Network. Incorporating MR physics constraints resulted in accurate T_1_-maps, M_0_-maps, and harmonized T_1_w images, producing stable brain segmentation masks regardless of the input domain. Our results demonstrated that using the Bloch equation as a constraint for deep neural network training can improve the accuracy of quantitative maps and harmonized T_1_w images even with a small number of datasets. We implemented the PhyCHarm framework using a supervised approach in this initial attempt. However, the framework could be used in an unsupervised manner or a self-supervised manner with large-scale datasets for next-generation harmonization. In future studies, PhyCHarm could be extended not only to different field strengths such as 1.5T or 7T, but also to other types of contrasts, such as T_2_w or FLAIR, as well as T_2_-maps.

## Data Availability

Dataset for the Quantitative Maps Generator: The dataset for the Quantitative Maps Generator in this paper is derived from the MICA-MICs dataset (Royer et al., 2022)
Dataset for the Harmonization Network (Traveling dataset): The dataset for the Harmonization Network in this paper is available on request from the corresponding author and can be shared following Institutional Review Board approval due to privacy or ethical restrictions.
Royer J, Rodriguez-Cruces R, Tavakol S, et al. An open MRI dataset for multiscale neuroscience. Sci Data. 2022;9(1):569. doi:10.1038/s41597-022-01682-y.
The source code is available at https://github.com/HUFS-AIIMST/PhyCHarm

https://portal.conp.ca/dataset?id=projects/mica-mics

## ACKNOWLEDGMENTS

Authors appreciate Megan Griffiths for editing the manuscript.

This work was supported by the National Research Foundation of Korea (NRF) grant funded by the Korean government (MSIT) (RS-2023-NR076864), by the MSIT (Ministry of Science, ICT), Korea, under the High-Potential Individuals Global Training Program (2021-0-01553), supervised by the IITP (Institute for Information & Communications Technology Planning & Evaluation), and the Hankuk University of Foreign Studies Research Fund.

## REFERENCES

1. Biberacher V, Schmidt P, Keshavan A, et al. Intra-and interscanner variability of magnetic resonance imaging based volumetry in multiple sclerosis. Neuroimage. 2016;142:188–197. doi:10.1016/j.neuroimage.2016.07.035.

2. Sampat MP, Healy BC, Meier DS, Dell’Oglio E, Liguori M, Guttmann CR. Disease modeling in multiple sclerosis: assessment and quantification of sources of variability in brain parenchymal fraction measurements. Neuroimage. 2010;52(4):1367–1373. doi:10.1016/j.neuroimage.2010.03.075.

3. Shinohara RT, Oh J, Nair G, et al; NAIMS Cooperative. Volumetric analysis from a harmonized multisite brain MRI study of a single subject with multiple sclerosis. AJNR Am J Neuroradiol. 2017;38(8):1501–1509. doi:10.3174/ajnr.A5254.

4. Bane O, Hectors SJ, Wagner M, et al. Accuracy, repeatability, and interplatform reproducibility of T1 quantification methods used for DCE-MRI: Results from a multicenter phantom study. Magn Reson Med. 2018;79(5):2564–2575. doi:10.1002/mrm.26903.

5. Hanson CA, Kamath A, Gottbrecht M, Ibrahim S, Salerno M. T2 relaxation times at cardiac MRI in healthy adults: A systematic review and meta-analysis. Radiology. 2020;297(2):344–351. doi:10.1148/radiol.2020200989.

6. Keenan KE, Gimbutas Z, Dienstfrey A, et al. Multi-site, multi-platform comparison of MRI T 1 measurement using the system phantom. PLoS One. 2021;16(6):e0252966. doi:10.1371/journal.pone.0252966.

7. Stikov N, Boudreau M, Levesque IR, Tardif CL, Barral JK, Pike GB. On the accuracy of T1 mapping: searching for common ground. Magn Reson Med. 2015;73(2):514–522. doi:10.1002/mrm.25135.

8. Karakuzu A, Biswas L, Cohen-Adad J, Stikov N. Vendor-neutral sequences and fully transparent workflows improve inter-vendor reproducibility of quantitative MRI. Magn Reson Med. 2022;88(3):1212–1228. doi:10.1002/mrm.29292.

9. Tustison NJ, Avants BB, Cook PA, et al. N4ITK: improved N3 bias correction. IEEE Trans Med Imaging. 2010;29(6):1310–1320. doi:10.1109/TMI.2010.2046908.

10. Beer JC, Tustison NJ, Cook PA, et al; Alzheimer’s Disease Neuroimaging Initiative. Longitudinal ComBat: A method for harmonizing longitudinal multi-scanner imaging data. Neuroimage. 2020;220:117129. doi:10.1016/j.neuroimage.2020.117129.

11. Choi KS. Deep learning applications in perfusion MRI: Recent advances and current challenges. Investig Magn Reson Imaging. 2022;26(4):246–255. doi:10.13104/imri.2022.26.4.246.

12. Fortin J-P, Parker D, Tunç B, et al. Harmonization of multi-site diffusion tensor imaging data. Neuroimage. 2017;161:149–170. doi:10.1016/j.neuroimage.2017.08.047.

13. Fortin J-P, Cullen N, Sheline YI, et al. Harmonization of cortical thickness measurements across scanners and sites. Neuroimage. 2018;167:104–120. doi:10.1016/j.neuroimage.2017.11.024.

14. Garcia-Dias R, Scarpazza C, Baecker L, et al. Neuroharmony: A new tool for harmonizing volumetric MRI data from unseen scanners. Neuroimage. 2020;220:117127. doi:10.1016/j.neuroimage.2020.117127.

15. Pomponio R, Erus G, Habes M, et al. Harmonization of large MRI datasets for the analysis of brain imaging patterns throughout the lifespan. Neuroimage. 2020;208:116450. doi:10.1016/j.neuroimage.2019.116450.

16. Bashyam VM, Doshi J, Erus G, et al; iSTAGING and PHENOM consortia. Deep generative medical image harmonization for improving cross-site generalization in deep learning predictors. J Magn Reson Imaging. 2022;55(3):908–916. doi:10.1002/jmri.27908.

17. Fatania K, Clark A, Frood R, et al. Harmonisation of scanner-dependent contrast variations in magnetic resonance imaging for radiation oncology, using style-blind auto-encoders. Phys Imaging Radiat Oncol. 2022;22:115–122. doi:10.1016/j.phro.2022.05.005.

18. Guan H, Liu Y, Yang E, Yap P-T, Shen D, Liu M. Multi-site MRI harmonization via attention-guided deep domain adaptation for brain disorder identification. Med Image Anal. 2021;71:102076. doi:10.1016/j.media.2021.102076.

19. Liu M, Maiti P, Thomopoulos S, et al. Style transfer using generative adversarial networks for multi-site MRI harmonization. Med Image Comput Comput Assist Interv. 2021;12903:313–322. doi:10.1007/978-3-030-87199-4_30.

20. Ren M, Dey N, Fishbaugh J, Gerig G. Segmentation-renormalized deep feature modulation for unpaired image harmonization. IEEE Trans Med Imaging. 2021;40(6):1519–1530. doi:10.1109/TMI.2021.3059726.

21. Robinson R, Dou Q, Coelho de Castro D, et al. Image-level harmonization of multi-site data using image-and-spatial transformer networks. Presented at: Medical Image Computing and Computer Assisted Intervention–MICCAI 2020: 23rd International Conference. Lima, Peru; October 4-8, 2020. p. 710–719. doi:10.1007/978-3-030-59728-3_69.

22. Yang Q, Li N, Zhao Z, Fan X, Chang EI-C, Xu Y. MRI cross-modality image-to-image translation. Sci Rep. 2020;10(1):3753. doi:10.1038/s41598-020-60520-6.

23. Zhang R, Pfister T, Li J. Harmonic unpaired image-to-image translation. Presented at: International Conference on Learning Representations (ICLR) 2019. New Orleans, LA; May 6-9, 2019. doi:10.48550/arXiv.1902.09727.

24. Ronneberger O, Fischer P, Brox T. U-net: Convolutional networks for biomedical image segmentation. In: Navab N, Hornegger J, Wells W, Frangi A, eds. Medical Image Computing and Computer-Assisted Intervention–MICCAI 2015. Cham, Switzerland: Springer; 2015. p. 234–241. doi:10.1007/978-3-319-24574-4_28.

25. Isola P, Zhu JY, Zhou T, Efros AA. Image-to-image translation with conditional adversarial networks. Presented at: 2017 IEEE Conference on Computer Vision and Pattern Recognition (CVPR). Honolulu, HI; 2017. p. 5967–5976. doi:10.1109/CVPR.2017.632.

26. Goodfellow I, Pouget-Abadie J, Mirza M, et al. Generative adversarial networks. Communications of the ACM. 2020;63(11):139–144. doi:10.1145/3422622.

27. Zhu JY, Park T, Isola P, Efros AA. Unpaired image-to-image translation using cycle-consistent adversarial networks. Presented at: 2017 IEEE International Conference on Computer Vision (ICCV). Venice, Italy; 2017. p. 2242–2251. doi:10.1109/ICCV.2017.244.

28. Kingma DP, Welling M. Auto-encoding variational bayes. arXiv:1312.6114; 2013. doi:10.48550/arXiv.1312.6114.

29. Choi Y, Choi M, Kim M, Ha JW, Kim S, Choo J. StarGAN: Unified Generative Adversarial Networks for Multi-domain Image-to-Image Translation. Presented at: 2018 IEEE/CVF Conference on Computer Vision and Pattern Recognition (CVPR). Salt Lake City, Utah; 2018. p. 8789–8797. doi:10.1109/CVPR.2018.00916.

30. Dewey BE, Zhao C, Reinhold JC, et al. DeepHarmony: A deep learning approach to contrast harmonization across scanner changes. Magn Reson Imaging. 2019;64:160–170. doi:10.1016/j.mri.2019.05.041.

31. Zuo L, Dewey BE, Liu Y, et al. Unsupervised MR harmonization by learning disentangled representations using information bottleneck theory. Neuroimage. 2021;243:118569. doi:10.1016/j.neuroimage.2021.118569.

32. Moyer D, Ver Steeg G, Tax CM, Thompson PM. Scanner invariant representations for diffusion MRI harmonization. Magn Reson Med. 2020;84(4):2174–2189. doi:10.1002/mrm.28243.

33. Torbati ME, Minhas DS, Laymon CM, et al. MISPEL: A supervised deep learning harmonization method for multi-scanner neuroimaging data. Med Image Anal. 2023;89:102926. doi:10.1016/j.media.2023.102926.

34. Wang Z, Bovik AC, Sheikh HR, Simoncelli EP. Image quality assessment: from error visibility to structural similarity. IEEE Trans Image Process. 2004;13(4):600–612. doi:10.1109/TIP.2003.819861.

35. Smith SM, Jenkinson M, Woolrich MW, et al. Advances in functional and structural MR image analysis and implementation as FSL. Neuroimage. 2004;23 Suppl 1:S208–S219. doi:10.1016/j.neuroimage.2004.07.051.

36. Royer J, Rodríguez-Cruces R, Tavakol S, et al. An open MRI dataset for multiscale neuroscience. Sci Data. 2022;9(1):569. doi:10.1038/s41597-022-01682-y.

37. Mugler JP 3rd, Brookeman JR. Three-dimensional magnetization-prepared rapid gradient-echo imaging (3D MP RAGE). Magn Reson Med. 1990;15(1):152–157. doi:10.1002/mrm.1910150117.

38. Marques JP, Kober T, Krueger G, van der Zwaag W, Van de Moortele P-F, Gruetter R. MP2RAGE, a self bias-field corrected sequence for improved segmentation and T1-mapping at high field. Neuroimage. 2010;49(2):1271–1281. doi:10.1016/j.neuroimage.2009.10.002.

39. Isensee F, Schell M, Pflueger I, et al. Automated brain extraction of multisequence MRI using artificial neural networks. Hum Brain Mapp. 2019;40(17):4952–4964. doi:10.1002/hbm.24750.

40. Tustison NJ, Cook PA, Holbrook AJ, et al. The ANTsX ecosystem for quantitative biological and medical imaging. Sci Rep. 2021;11(1):9068. doi:10.1038/s41598-021-87564-6.

41. Kingma DP, Ba J. Adam: A method for stochastic optimization. Presented at: International Conference for Learning Representations (ICLR) 2015. San Diego, CA; May 7-9, 2015. doi:10.48550/arXiv.1412.6980.

42. Azizi S, Culp L, Freyberg J, et al. Robust and data-efficient generalization of self-supervised machine learning for diagnostic imaging. Nat Biomed Eng. 2023;7(6):756–779. doi:10.1038/s41551-023-01049-7.

