## supplementary material for "A preliminary attempt to harmonize using physics-constrained deep neural networks for multisite and multiscanner MRI datasets (PhyCHarm)"

TABLE S1 Details of the Harmonization Network dataset (traveling dataset).

| **Scan parameter** | **Siemens Trio  (Site 1)** | **GE SIGNA  (Site 2)** | **Philips Ingenia CS  (Site 3)** |
| --- | --- | --- | --- |
| Field strength | 3T | 3T | 3T |
| Echo time (ms) | 2.12 | 3.192 | 3.271 |
| Repetition time (ms) | 2400 | 458 | - |
| Inversion time (ms) | 1000 | 450 | - |
| Echo train length | - | - | 192 |
| Matrix size | 224 × 256 × 256 | 192 × 256 × 256 | 160 × 768 × 768 |
| Resolution (mm^3^) | 1 × 1 × 1 | 1 × 1 × 1 | 1 × 0.33 × 0.33 |
| Number of subjects | 9 | 9 | 9 |

TABLE S2 Quantitative evaluation scores for harmonization methods (validation).

| **Harmonization method** | **GE → Siemens** | **Philips → Siemens** | **Siemens → GE** | **Philips → GE** | **Siemens → Philips** | **GE → Philips** |
| --- | --- | --- | --- | --- | --- | --- |
|  | **SSIM** | | | | | |
| Without harmonization | 0.7914 | 0.8173 | 0.7914 | 0.9226 | 0.8173 | 0.9226 |
| U-Net | 0.9666 | 0.9646 | 0.9673 | 0.9671 | 0.9747 | 0.9722 |
| Pix2Pix | 0.9663 | 0.9555 | 0.9660 | 0.9648 | 0.9702 | 0.9727 |
| CALAMITI | 0.9408 | 0.9404 | 0.9386 | 0.9341 | 0.9369 | 0.9360 |
| PhyCHarm | **0.9789** | **0.9778** | **0.9718** | **0.9888** | **0.9784** | **0.9768** |
|  | **PSNR** | | | | | |
| Without harmonization | 21.8551 | 25.7831 | 21.8551 | 25.2566 | 25.7831 | 25.2566 |
| U-Net | 28.0568 | 27.0684 | 28.4797 | 26.6743 | 28.5396 | 28.6781 |
| Pix2Pix | 28.4520 | 25.2289 | 26.8872 | 26.4553 | 27.1151 | 28.9024 |
| CALAMITI | 28.9575 | 27.9198 | 28.3207 | 27.0025 | 28.2772 | 26.6447 |
| PhyCHarm | **32.2353** | **31.4737** | **32.7711** | **36.4058** | **33.3574** | **33.1869** |

*Note:* → denotes harmonizing the input domain to the target domain. The bold numbers represent the highest SSIM and PSNR values. SSIM and PSNR for the method without harmonization were calculated between the original data from the target domain and the original data from the input domain. SSIM and PSNR for the method with harmonization were measured between the original data from the target domain and the harmonized data from the input domain. SSIM = structural similarity index measure; PSNR = peak-signal-to-noise ratio.

TABLE S3 Dice scores (validation).

| **Harmonization method** | **GE → Siemens** | **Philips → Siemens** | **Siemens → GE** | **Philips → GE** | **Siemens → Philips** | **GE → Philips** |
| --- | --- | --- | --- | --- | --- | --- |
|  | **GM** | | | | | |
| Without harmonization | 0.8593 | 0.8747 | 0.8593 | 0.8783 | 0.8747 | 0.8783 |
| U-Net | 0.8829  (*p* < 0.05) | 0.8690  (*p* = 0.108) | 0.8481  (*p* < 0.05) | 0.8610  (*p* < 0.05) | 0.8734  (*p* = 0.276) | 0.8791  (*p* = 0.905) |
| Pix2Pix | 0.8700  (*p* < 0.05) | 0.8417  (*p* < 0.05) | 0.8258  (*p* < 0.05) | 0.8299  (*p* < 0.05) | 0.8531  (*p* < 0.05) | 0.8692  (*p* < 0.05) |
| CALAMITI | 0.7339  (*p* < 0.05) | 0.7795  (*p* < 0.05) | 0.7442  (*p* < 0.05) | 0.7737  (*p* < 0.05) | 0.7581  (*p* < 0.05) | 0.7649  (*p* < 0.05) |
| PhyCHarm | **0.8977^*^**  **(*p* < 0.05)** | **0.8909^*^**  **(*p* < 0.05)** | **0.8638^*^**  **(*p* < 0.05)** | **0.8972^*^**  **(*p* < 0.05)** | **0.8923^*^**  **(*p* < 0.05)** | **0.8866^*^**  **(*p* < 0.05)** |
|  | **WM** | | | | | |
| Without harmonization | 0.9136 | 0.9140 | 0.9136 | 0.9203 | 0.9140 | 0.9203 |
| U-Net | 0.9257  (*p* < 0.05) | 0.9208  (*p* < 0.05) | 0.9096  (*p* < 0.05) | 0.9187  (*p* < 0.05) | 0.9026  (*p* < 0.05) | 0.9208  (*p* < 0.05) |
| Pix2Pix | 0.9182  (*p* = 0.580) | 0.9008  (*p* < 0.05) | 0.8974  (*p* < 0.05) | 0.9014  (*p* < 0.05) | 0.8927  (*p* < 0.05) | 0.9195  (*p* < 0.05) |
| CALAMITI | 0.8576  (*p* < 0.05) | 0.8741  (*p* < 0.05) | 0.8489  (*p* < 0.05) | 0.8770  (*p* < 0.05) | 0.8442  (*p* < 0.05) | 0.8580  (*p* < 0.05) |
| PhyCHarm | **0.9390^*^**  **(*p* < 0.05)** | **0.9332^*^**  **(*p* < 0.05)** | **0.9141**  **(*p* = 0.393)** | **0.9357^*^**  **(*p* < 0.05)** | **0.9171**  **(*p* = 0.431)** | **0.9213^*^**  **(*p* < 0.05)** |

*Note:* Dice scores are displayed as mean values. Dice scores for the method without harmonization were calculated between the original data from the target domain and the original data from the input domain. Dice scores for the method with harmonization were calculated between the original data from the target domain and the harmonized data from the input domain. A Wilcoxon signed-rank test was performed to compare the Dice scores for the method without harmonization with those for the other harmonization methods. Scores marked with * indicate significantly higher Dice scores compared to the method without harmonization (*p* < 0.05). The bold values denote the highest Dice scores. GM = gray matter; WM = white matter

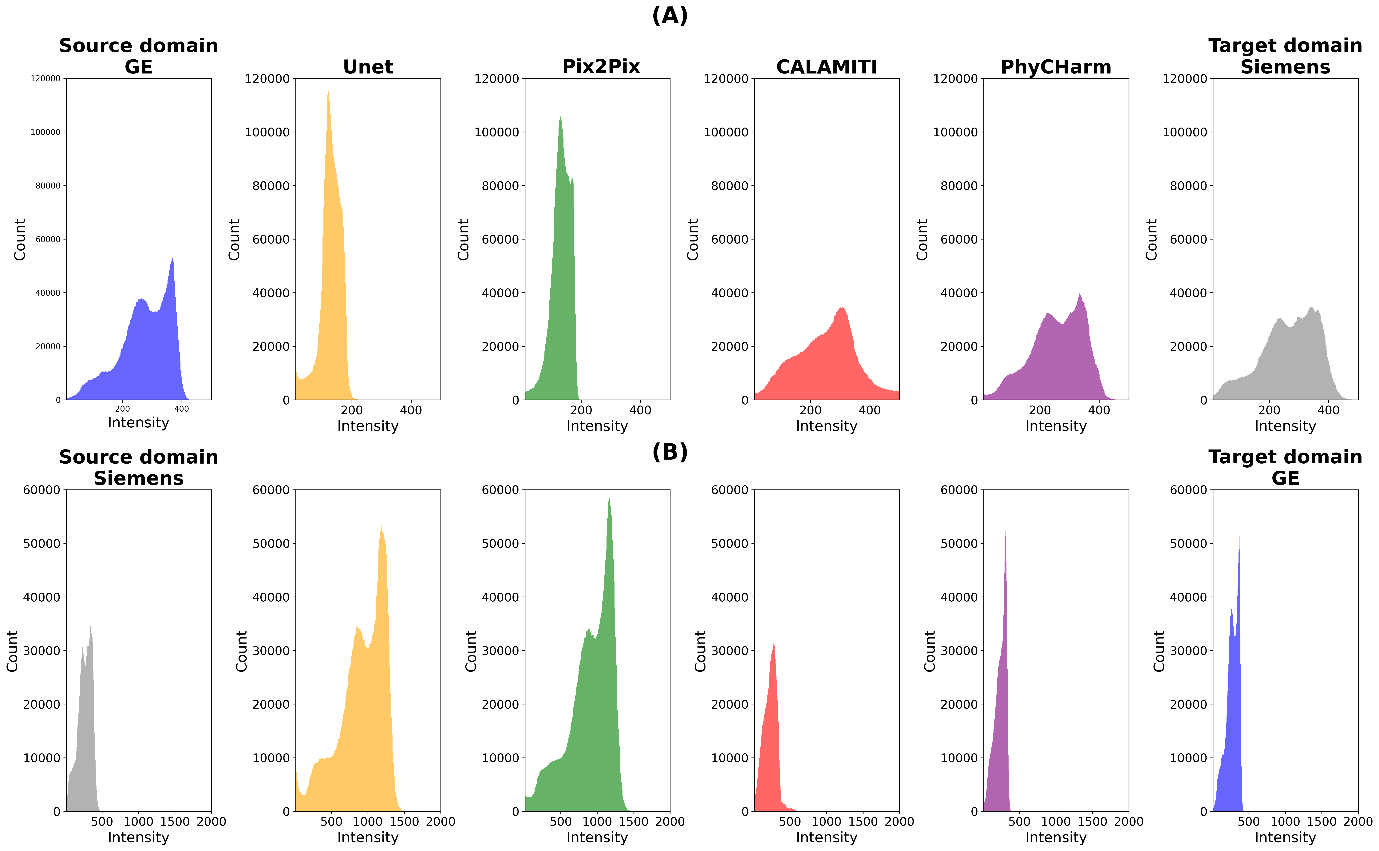

FIGURE S1 Comparison of histograms for GE and Siemens harmonization. The original datasets are represented in blue (GE) and gray (Siemens). (A) The case of GE to Siemens harmonization. The histograms for the U-Net and Pix2Pix results show a notable difference in intensity range and histogram shape compared to those of the target domain. In contrast, the histograms for CALAMITI and PhyCHarm closely resemble those of the target domain. In the intensity range of 200 to 300, the peak observed in the target domain histogram does not appear in CALAMITI but is visible in PhyCHarm. (B) The case of Siemens to GE harmonization. The intensity ranges of U-Net and Pix2Pix do not align with those of the target domain. Although CALAMITI demonstrates a histogram shape comparable to that of the target domain, the highest peak observed in the target domain histogram is not shown in CALAMITI. PhyCHarm demonstrates the histogram shape closest to that of the target domain; however, the second-largest peak in the target domain is missing from the PhyCHarm histogram.

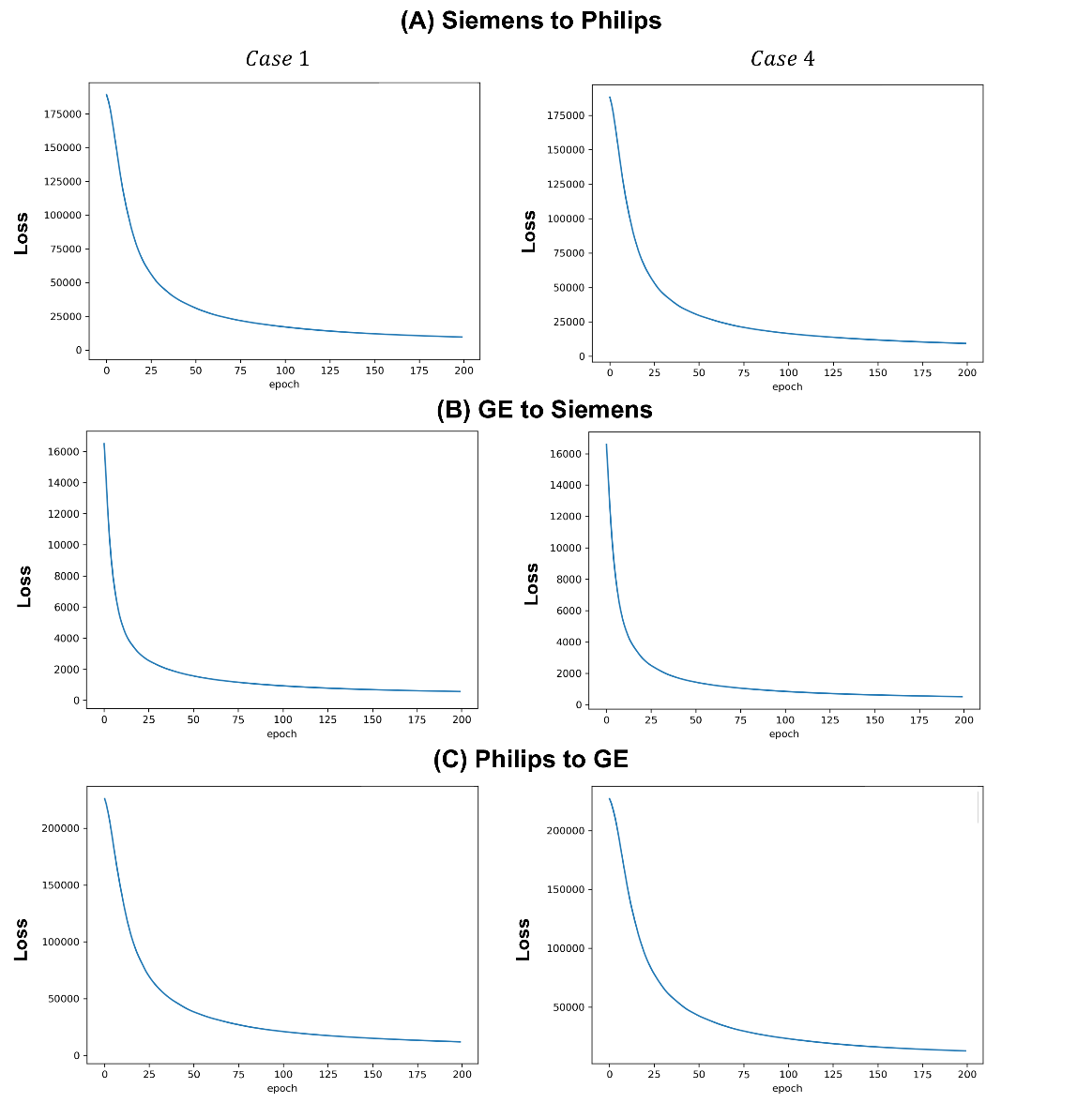

**FIGURE S2** Comparison of the training loss progression of the Harmonization Network. The loss reduction varies depending on the weight values for the consistency loss ($\lambda_{3}$) from the Quantitative Maps Generator. (Case 1: $\lambda_{3}$= 0; Case 4: $\lambda_{3}$= ${1e}^{-6}$). Case 4 shows rapid loss reduction compared to Case 1, as shown in (A) Siemens to Philips, (B) GE to Siemens, and (C) Philips to GE harmonization.

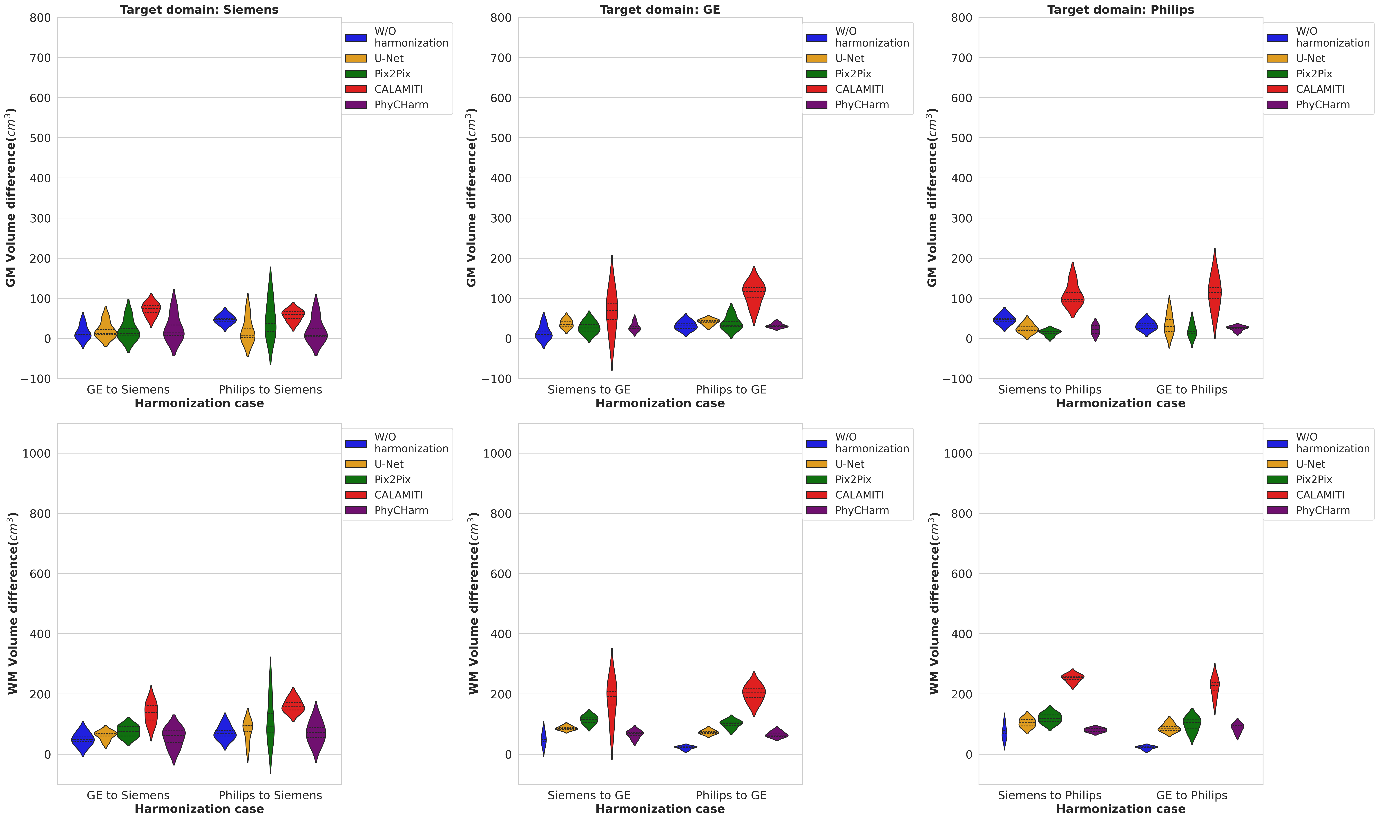

FIGURE S3 Gray matter (GM) and white matter (WM) volumetric differences in the validation results. The first row shows the GM volumetric differences; the second row shows the WM volumetric differences. The volumetric difference was calculated between the input domain and target domain volumes. “W/O harmonization” represents the results obtained without applying harmonization. For GM with Siemens as the target domain, U-Net, Pix2Pix, and PhyCHarm demonstrate reduced volumetric differences but high variance, whereas CALAMITI shows increased volumetric differences. When the target domain is GE, PhyCharm shows the smallest volumetric difference and lower variance than that seen with U-Net, Pix2Pix, and CALAMITI. When the target domain is Philips, Pix2Pix and PhyCharm show reduced volumetric differences compared to the method without harmonization. For WM, none of the harmonization methods enhance the volumetric differences compared to the method without harmonization.
